# Characterisation of the mechanism by which a nonsense variant in *RYR2* results in ventricular arrhythmia

**DOI:** 10.1101/2021.03.16.21252576

**Authors:** Claire Hopton, Anke J Tijsen, Leonid Maizels, Gil Arbel, Amira Gepstein, Nicola Bates, Benjamin Brown, Irit Huber, Susan J Kimber, William G Newman, Luigi Venetucci, Lior Gepstein

**Affiliations:** Division of Evolution and Genomic Sciences, Faculty of Biology, Medicine and Health, University of Manchester, Manchester, UK.; Manchester Centre for Genomic Medicine, Manchester University NHS Foundation Trust, Health Innovation Manchester, Manchester, UK.; Division of Cardiovascular Sciences, Faculty of Biology, Medicine and Health, University of Manchester, Manchester, UK.; The Rappaport Faculty of Medicine and Research Institute, Technion-Institute of Technology, Haifa, Israel.; Amsterdam UMC, University of Amsterdam, Department of Experimental Cardiology, Amsterdam Cardiovascular Sciences, Meibergdreef 9, Amsterdam, The Netherlands.; Division of Cardiology, Sheba Medical Center Hospital, Tel Hashomer, Israel.; Division of Cell Matrix Biology and Regenerative Medicine, Faculty of Biology, Medicine and Health, University of Manchester, Manchester, UK.; Department of Cardiology, Wythenshawe Hospital, Manchester University NHS Foundation Trust, Manchester, UK.; Manchester Heart Centre, Manchester University NHS Foundation Trust, Health Innovation Manchester, Manchester, UK.; Cardiology Department, Rambam Health Care Campus, Haifa, Israel.

**Keywords:** RYR2, CPVT, ventricular arrhythmia, induced pluripotent stem cells, nonsense variant

## Abstract

**Background:** Heterozygous variants in the cardiac ryanodine receptor gene (*RYR2*) cause catecholaminergic polymorphic ventricular tachycardia (CPVT). Most pathogenic *RYR2* variants are missense variants which result in a gain of function, causing ryanodine receptors to be increasingly sensitive to activation by calcium, have an increased open probability and an increased propensity to develop calcium waves. However, some *RYR2* variants can lead to arrhythmias by a loss of function mechanism.

**Objective:** To understand the mechanism by which a novel nonsense variant in *RYR2* p.(Arg4790Ter) leads to ventricular arrhythmias.

**Methods:** Human induced pluripotent stem cells (hiPSCs) harbouring the novel nonsense variant in *RYR2* were differentiated into cardiomyocytes (RYR2-hiPSC-CMs) and molecular and calcium handling properties were studied.

**Results:** RYR2-hiPSC-CMs displayed significant calcium handling abnormalities at baseline and following treatment with isoproterenol. Treatment with carvedilol and nebivolol resulted in a significant reduction in calcium handling abnormalities in the RYR2-hiPSC-CMs. Expression of the mutant *RYR2* allele was confirmed at the mRNA level and partial silencing of the mutant allele resulted in a reduction in calcium handling abnormalities at baseline.

**Conclusion:** The nonsense variant behaves similarly to other gain of function variants in *RYR2*. Carvedilol and nebivolol may be suitable treatments for patients with gain of function *RYR2* variants.

## Introduction

The *RYR2* gene encodes for the cardiac ryanodine receptor, a major calcium handling channel located within cardiomyocytes. Most disease-causing variants in *RYR2* are missense variants and result in an autosomal dominant form of catecholaminergic polymorphic ventricular tachycardia (CPVT1, MIM 604772). CPVT is characterised by adrenergically mediated arrhythmias, typically bidirectional or polymorphic VT, in the presence of a structurally normal heart. Most *RYR2* variants which have been functionally studied have been shown to result in a gain of function of RYR2, causing an increased sensitivity of the channel to activation by calcium and an increased open probability. Despite this, a small number of missense variants in *RYR2* have been shown to result in a loss of function and possibly cause arrhythmias by a different mechanism however further work is needed to fully understand these mechanisms (1). Determining the mechanisms by which *RYR2* variants can lead to arrhythmias is important as treatment efficacy may be dependent on the underlying mechanism which leads to arrhythmias.

We identified a novel nonsense variant in *RYR2* in a young woman who suffered a cardiac arrest. To understand whether this variant causes arrhythmias by a different mechanism to the majority of missense variants in *RYR2*, we studied cardiomyocytes differentiated from hiPSCs generated from the woman.

## Methods

For detailed methods see Supplementary Materials.

### Study approval

Ethical approval for this study was obtained from the Central Manchester Research Ethics Committee (11/H1003/3). Consent was obtained from all participants involved in the study.

### Generation and maintenance of human induced pluripotent stem cells and differentiation into cardiomyocytes

hiPSCs were generated by retroviral infection of dermal fibroblasts obtained from a young woman carrying a heterozygous nonsense variant in *RYR2*, p.(Arg4790Ter).

hiPSCs were cultured on 1:200 growth-factor reduced Matrigel (Corning) in mTeSR1 culture medium (Stem Cell Technologies) with the medium being refreshed daily. Cardiomyocytes were generated from a monolayer of hiPSCs using a chemically defined previously published differentiation protocol (2).

### Quantitative reverse transcriptase polymerase reaction (RT-PCR) and western blot

Total RNA and protein were extracted from nonsense variant carrying hiPSC-CMs (RYR2-hiPSC-CMs) and control hiPSC-CMs. Allele-specific RT-PCR was performed using primers designed on single nucleotide polymorphisms (SNPs) in *RYR2* (see Supplementary Materials, Table S2).

Western blot analysis was performed to assess total RYR2 protein levels and determine whether the mutant RYR2 protein was present in RYR2-hiPSC-CMs. Two RYR2 antibodies were used; ARP106/1 (gift from Prof William’s laboratory Swansea, 1:500) reacts to an epitope in the far C-terminus (aa 4957-4967) which lies distal to the p.(Arg4790Ter) variant and sc-376507 RYR2 antibody (Santa Cruz, 1:500) which reacts to an epitope in the N-terminus, a region common to both the mutant and wild-type RYR2. A ratio of expression of these was then calculated for the RYR2-hiPSC-CMs and compared to the expression ratio in the control hiPSC-CMs.

### Laser Confocal Ca2+ imaging

hiPSC-CMs were enzymatically dissociated with TrypLE Express (Life Technologies) and plated onto Matrigel coated glass bottom optical culture dishes (MatTek Coorporation). Cells were loaded with 5µM Fluo-4AM (Molecular Probes). Calcium transients were recorded from spontaneously beating single hiPSC-CMs using the line scan mode of a Zeiss LSM-710 or LSM7 confocal system. All experiments were performed in tyrodes solution at 37°C. hiPSC-CMs were line scanned at baseline, incubated with 10µM isoproterenol for 20 minutes and then re-scanned.

Store overload induced calcium release (SOICR) experiments were undertaken as previously described by Itzhaki et al (3). In summary, hiPSC-CMs were treated with tetrodotoxin (10µM), lidocaine (50µM) and caesium chloride (5mM). hiPSC-CMs were then exposed to external solutions of varying calcium concentrations. Cells were scanned for 60 seconds using the line scan mode and spontaneous calcium releases were recorded.

For beta-blocker experiments, hiPSC-CMs were line scanned at baseline, incubated with a beta-blocker for 20 minutes and then re-scanned.

### shRNA design and generation

Allele-specific short hairpin RNAs (shRNAs) to target the mutant *RYR2* allele were designed. The allele-specificity was based on the nonsense variant and in each shRNA the mutation recognition site was located in a different position (Supplementary Materials, Table S3).

RYR2-hiPSC-CMs were lentivirally transduced with the shRNAs and RNA was subsequently extracted to perform allele-specific RT-PCR to assess the expression of the mutant and wild-type alleles.

For calcium imaging, the shRNAs were cloned into a vector containing Ds-Red, in order to identify effectively transduced cells. The transduced hiPSC-CMs were loaded with Fluo-4-AM. Cells which had been successfully transduced, identified by DsRed, were line scanned.

## Results

### Clinical Details

The novel nonsense variant in *RYR2*, c.14368C>T, p.(Arg4790Ter), was identified in a woman in her 30’s who suffered a cardiac arrest. Investigations following the cardiac arrest revealed an unremarkable resting ECG with no evidence of a prolonged QT interval or a Brugada pattern. A cardiac MRI revealed no structural cardiac abnormalities and left ventricular systolic function was reported as within normal limits. A coronary angiogram was normal. A dual chamber ICD was implanted after her cardiac arrest and the patient is currently on bisoprolol (2.5mg BD). Treatment with beta-blockers has fully controlled the patient’s symptoms and no further arrhythmic episodes have been documented since her cardiac arrest.

The proband initially underwent genetic testing using Next Generation Sequencing of a panel of 57 genes associated with inherited cardiac conditions (Manchester Regional Genetics Laboratory) and subsequently had whole exome sequencing (see Supplementary Materials) which identified the p.(Arg4790Ter) *RYR2* variant but no other likely pathogenic variants. The p.(Arg4790Ter) variant is not listed on dbSNP or gnomAD (n>140,000) (4) as a rare polymorphism. Based on the ACMG guidelines the p.(Arg4790Ter) variant would be classified as a variant of uncertain significance (5), however most variants of *RYR2* may fall into this category as they are novel or very rare (6). It is important to note that despite the patient suffering episodes of syncope associated with emotional stress and a cardiac arrest, polymorphic VT has never been documented which makes the phenotype of the patient atypical.

### Characterisation of calcium handling in the RYR2-hiPSC-CMs

Patient-specific hiPSCs harbouring the p.(Arg4790Ter) variant were generated. The presence of the nonsense variant was confirmed by sequencing gDNA (Supplementary Data, Figure S1). The pluripotency of the hiPSCs was confirmed and the cells were successfully differentiated into spontaneously beating cardiomyocytes, RYR2-hiPSC-CMs (Supplementary Data, Figures S3, S4 and S5).

Fluorescent calcium imaging revealed marked intracellular calcium transient abnormalities in the RYR2-hiPSC-CMs. These abnormalities were mainly large double and triple humped calcium transients (Figure 1A and B), however some were broader multiple peaked transients in which the peaks appeared less deep and narrower (A(ii)). The percentage of cells displaying calcium handling abnormalities was significantly higher in the hiPSC-CMs derived from the two patient-specific RYR2 hiPSC clones (70.9% and 64.9%) as compared to healthy-control cells (28.9%, p<0.001) (Figure 1B). Moreover, although calcium handling abnormalities were observed in some of the control cells, these tended to be less complex than those seen in the RYR2-hiPSC-CMs. There was no significant difference in the proportion of cells displaying abnormalities between cardiomyocytes generated from the two RYR2-hiPSC clones.

**Figure 1.**
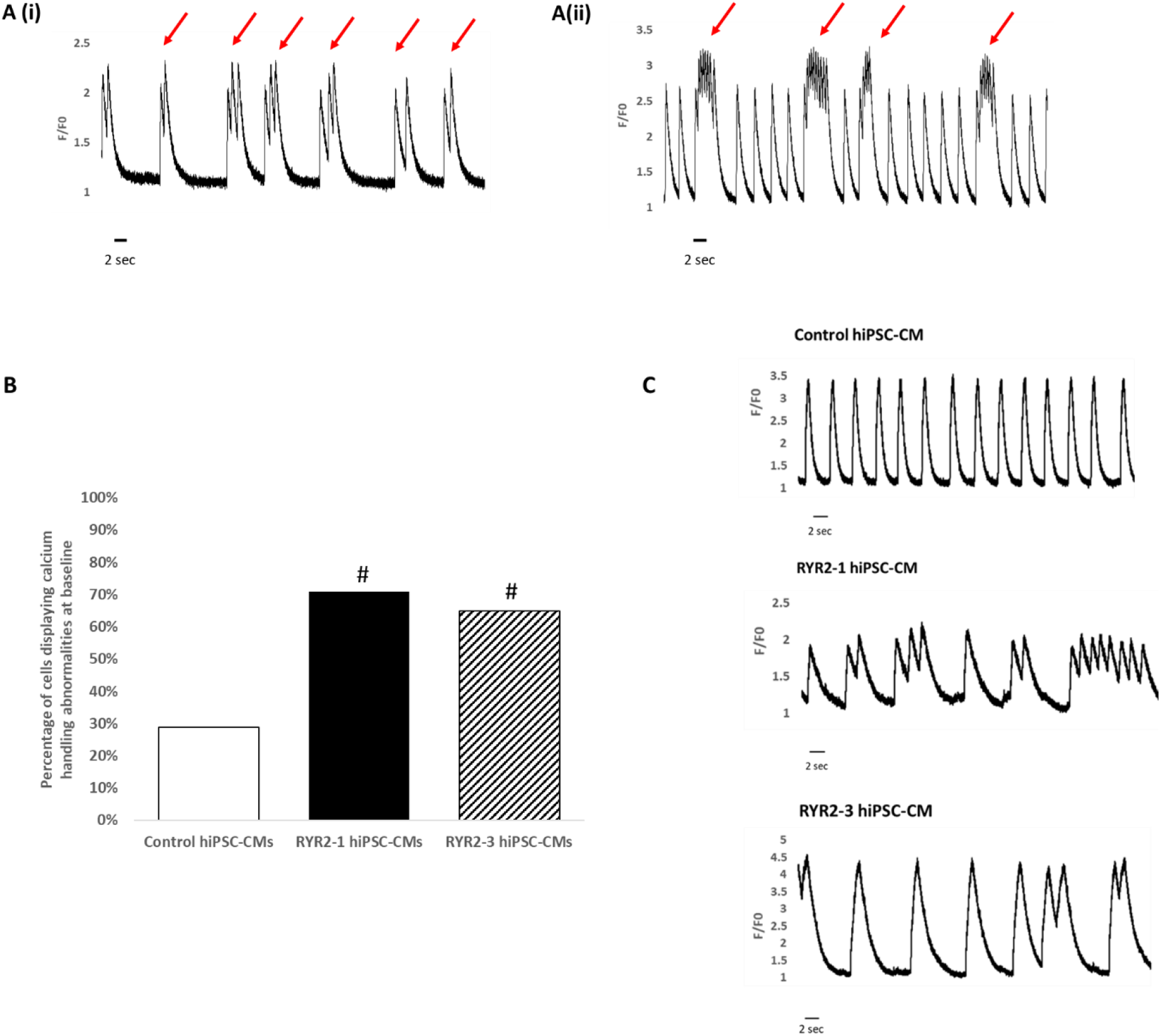
Calcium handling abnormalities in patient-specific RYR2-hiPSC-CMs at baseline. (A) Line scan image showing changes in intracellular calcium in single RYR2-hiPSC-CMs at baseline illustrating the differences between double and triple peaked and broader multiple peaked transients. (Ai) note the double and triple peaked transients (red arrows). (Aii) Note the broad multiple peaked transients (red arrows). (B) hiPSC-CMs generated from both RYR2 clones (RYR2-1 and RYR2-3) displayed significantly more calcium handling abnormalities compared to healthy-control hiPSC-CMs (control 28.9% n=121, RYR2-1 70.9% n=103, RYR2-3 64.9% n=259), # p<0.001. (C) Line scan image showing changes in intracellular calcium in single hiPSC-CMs at baseline.

#### Adrenergic stimulation

As variants in *RYR2* are associated with adrenergically mediated arrhythmias, we tested the effect of adrenergic stimulation on the hiPSC-CMs using isoproterenol. Focusing only on the hiPSC-CMs that displayed normal calcium transients at baseline, we noted that 52.6% (10 of 19) of the RYR2-hiPSC-CMs developed calcium transient abnormalities after the application of 10µM isoproterenol, significantly more than seen in the healthy-control cells (19.0%, n=21, p<0.05). Cells typically developed multiple peaked transients after being incubated with isoproterenol (Figure 2).

**Figure 2.**
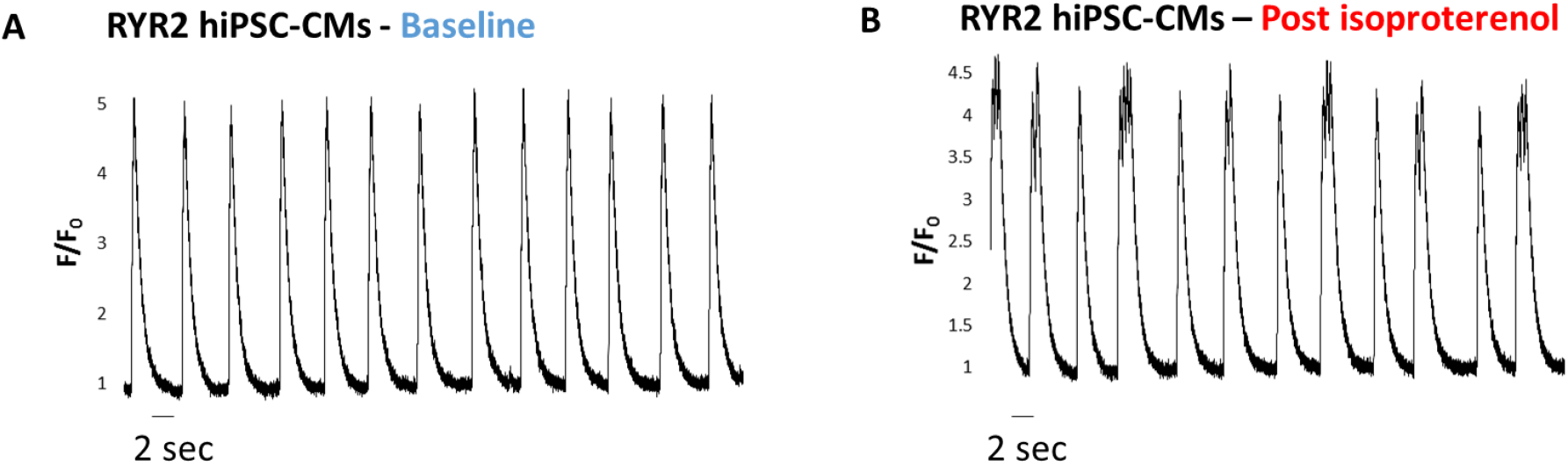
Whole cell [Ca2+]i transients in a RYR2-hiPSC-CM at baseline (A) and after treatment with isoproterenol (B). Note the development of double and triple humped transients after treatment with isoproterenol.

#### Store overload-induced calcium release

The p.(Arg4790Ter) variant is predicted to result in a truncated protein with loss of a region which has been implicated in luminal calcium sensing (7). Increasing the sensitivity of ryanodine receptors to luminal calcium facilitates the development of spontaneous calcium release. We therefore sought to assess whether the p.(Arg4790Ter) variant increased the propensity to develop spontaneous calcium releases at varying external calcium concentrations, the store overload-induced calcium release (SOICR) threshold. As demonstrated in Figure 3, SOICR events were noted in both RYR2- and healthy-control hiPSC-CMs, and their occurrence increased with elevated calcium concentration. Importantly, SOICR threshold was significantly reduced in the RYR2-hiPSC-CMs and the percentage of oscillating RYR2-hiPSC-CMs was significantly higher than of control hiPSC-CMs for each calcium concentration (p<0.001 for all calcium concentrations, n=29 for RYR2 and n=36 for control, Figure 3).

**Figure 3.**
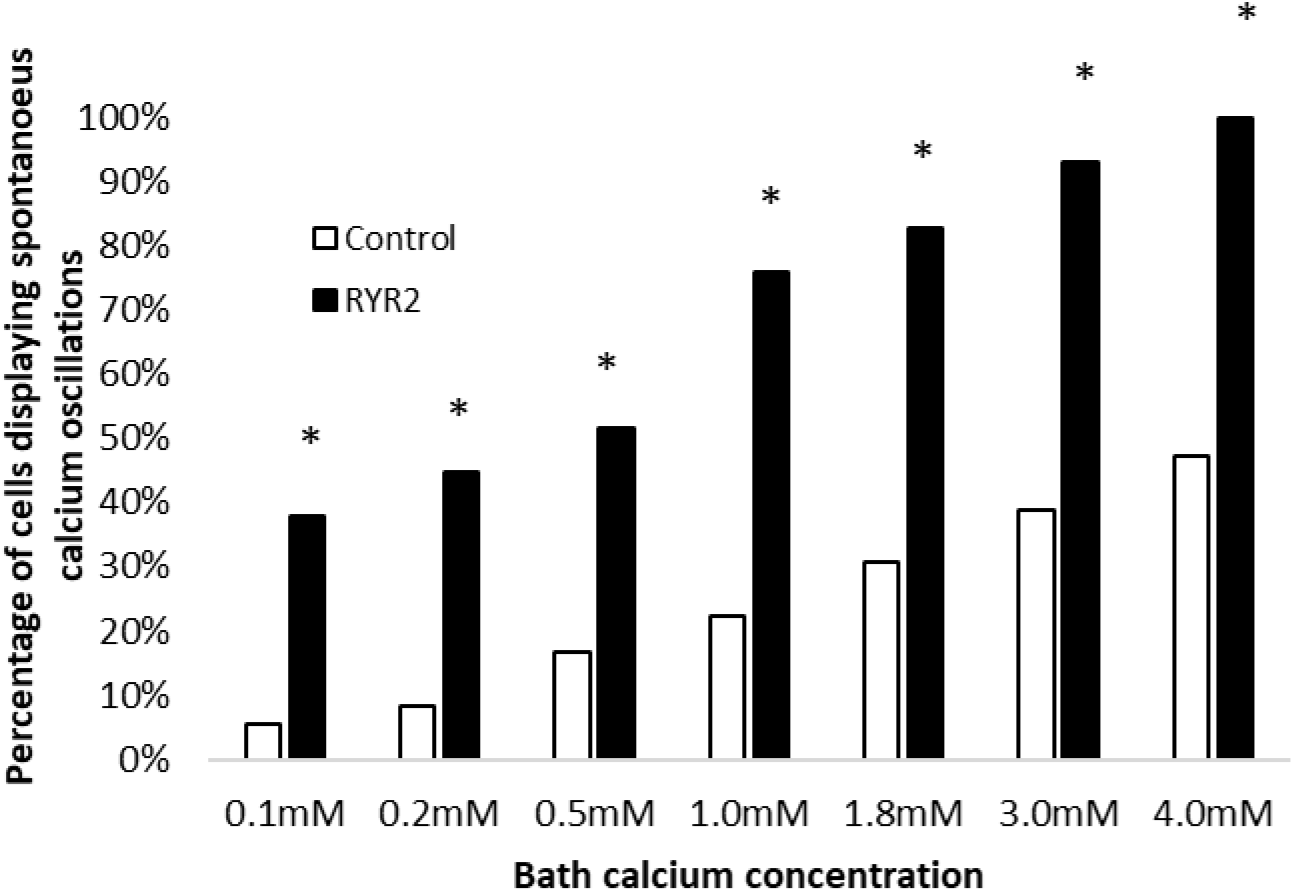
Percentage of RYR2-hiPSC-CMs (black) and healthy-control hiPSC-CMs (white) displaying spontaneous calcium oscillations when bathed in solutions with varying calcium concentrations. n= 29 and n=36 for RYR2 and control hiPSC-CMs respectively, * p<0.001 for all bath calcium concentrations.

These experiments clearly demonstrate that the p.(Arg4790Ter) variant increases the propensity of hiPSC-CMs to develop spontaneous calcium releases and suggests that the nonsense variant causes a gain of function phenotype.

### Drug screening in the RYR2-hiPSC-CMs

Both carvedilol and nebivolol inhibit ryanodine receptors. To confirm that the abnormalities we detected in the RYR2-hiPSC-CMs are caused by a gain of function we tested the effects of these two beta-blockers on the RYR2-hiPSC-CMs. To ensure the effects observed were not due to the beta-blocker properties of these drugs, we performed these experiments in the absence of adrenergic stimulation and tested the effects of several other beta-blockers as control. Treatment with both nebivolol and carvedilol resulted in significant reductions in the proportion of cells displaying calcium handling abnormalities (Figure 4). Treatment with labetalol also resulted in a small reduction in the proportion of hiPSC-CMs displaying abnormal calcium handling, however not to the magnitude seen with nebivolol or carvedilol. These findings confirm that the p.(Arg4790Ter) variant leads to a gain of function.

**Figure 4.**
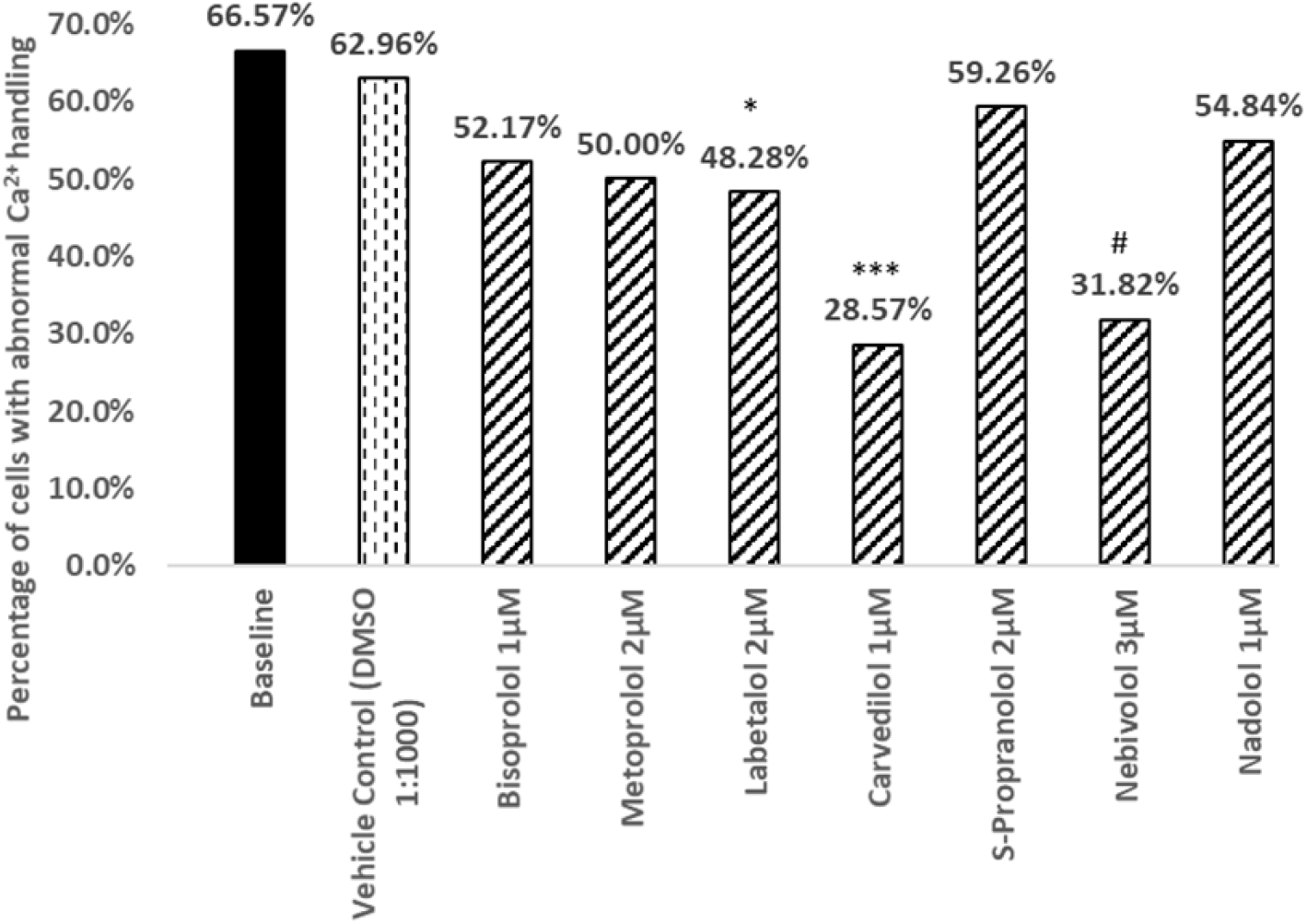
Effect of different beta-blockers on the proportion of RYR2-hiPSC-CMs displaying calcium handling abnormalities. Treatment with 2µM labetalol, 1µM carvedilol and 1µM nebivolol led to significant reductions in the proportion of cells displaying calcium transient abnormalities (*p<0.05, *** p<0.005, # p<0.001 respectively). Treatment with the other beta-blockers tested resulted in no significant difference. Baseline n=362, vehicle control n=27, bisoprolol n=23, metoprolol n=28, labetalol n=29, carvedilol n=14, S-propranolol n=27, nebivolol n=22, nadolol n=31.

### The effect of the p.(Arg4790Ter) variant on the presence of the RYR2 protein

To understand how the p.(Arg4790Ter) variant leads to a gain of function we designed an allele-specific RT-PCR based on a SNP (rs684923, c.7806C>T, MAF 45%) in *RYR2*, aiming to determine whether the mutant *RYR2* allele is expressed. Sequencing of gDNA confirmed that the RYR2-hiPSC-CMs were heterozygous for the SNP rs684923 whilst the control cells were homozygous (Figure 5A). The allele-specific RT-PCR showed expression of both alleles at the mRNA level in hiPSC-CMs derived from two different RYR2-hiPSC clones (RYR2-1 and RYR2-3) (Figure 5B). This finding suggests that the transcript escapes nonsense mediated decay. As expected, RT-PCR only detected one allele in the control hiPSC-CMs confirming the specificity of the primers. Despite expression of both alleles in the RYR2-hiPSC-CMs, total *RYR2* expression was found to be significantly reduced in the RYR2-hiPSC-CMs compared to expression in control (UN1-22) hiPSC-CMs and also hiPSC-CMs generated from another independent control line (Figure 5C). There was no significant difference in total RYR2 expression between hiPSC-CMs derived from the two different healthy-control lines.

**Figure 5.**
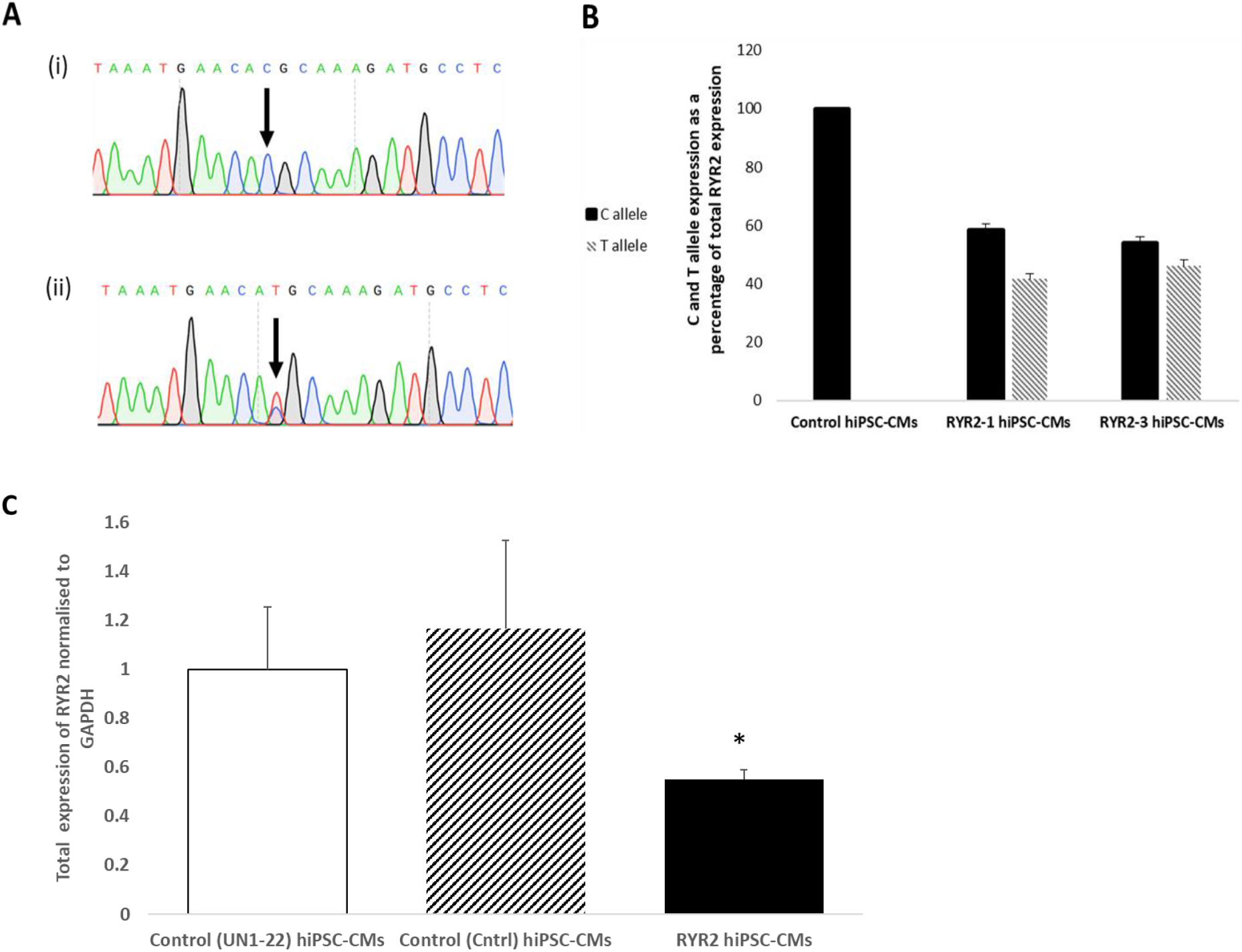
RYR2 expression in hiPSC-CMs harbouring the nonsense variant. **(A)** Sequencing of DNA extracted from healthy-control hiPSCs showed that the line is homozygous for the rs684923 SNP in *RYR2* (A(i)) whilst the RYR2 line is heterozygous for the SNP (A(ii)). (B) Allele-specific RT-PCR confirmed the expression of both RYR2 alleles in hiPSC-CMs generated from both clones of the RYR2 line (RYR2-1 and RYR2-3). Only the C allele was detected in control hiPSC-CMs confirming the specificity of the primers. (C) RT-PCR for total *RYR2* expression performed on RNA extracted from hiPSC-CMs derived from two different control lines (UN1-22 and Ctrl) and the RYR2-hiPSC-CMs (n=4, 2 and 5 respectively). In RYR2-hiPSC-CMs, total *RYR2* expression was significantly reduced compared to expression in the control lines (* p<0.05). There was no significant difference in *RYR2* expression between the two healthy-control lines.

Western blotting showed a significant reduction in total RYR2 protein levels in the RYR2-hiPSC-CMs compared to control (Figure 6B). There was no significant difference in the ratio of expression of the N-terminus to C-terminus in the RYR2-hiPSC-CMs compared to control (Figure 6D). An elevated N-terminus to C-terminus ratio in the RYR2-hiPSC-CMs compared to control would be expected if the truncated protein is present. These data suggest that the truncated RYR2 is not present.

**Figure 6.**
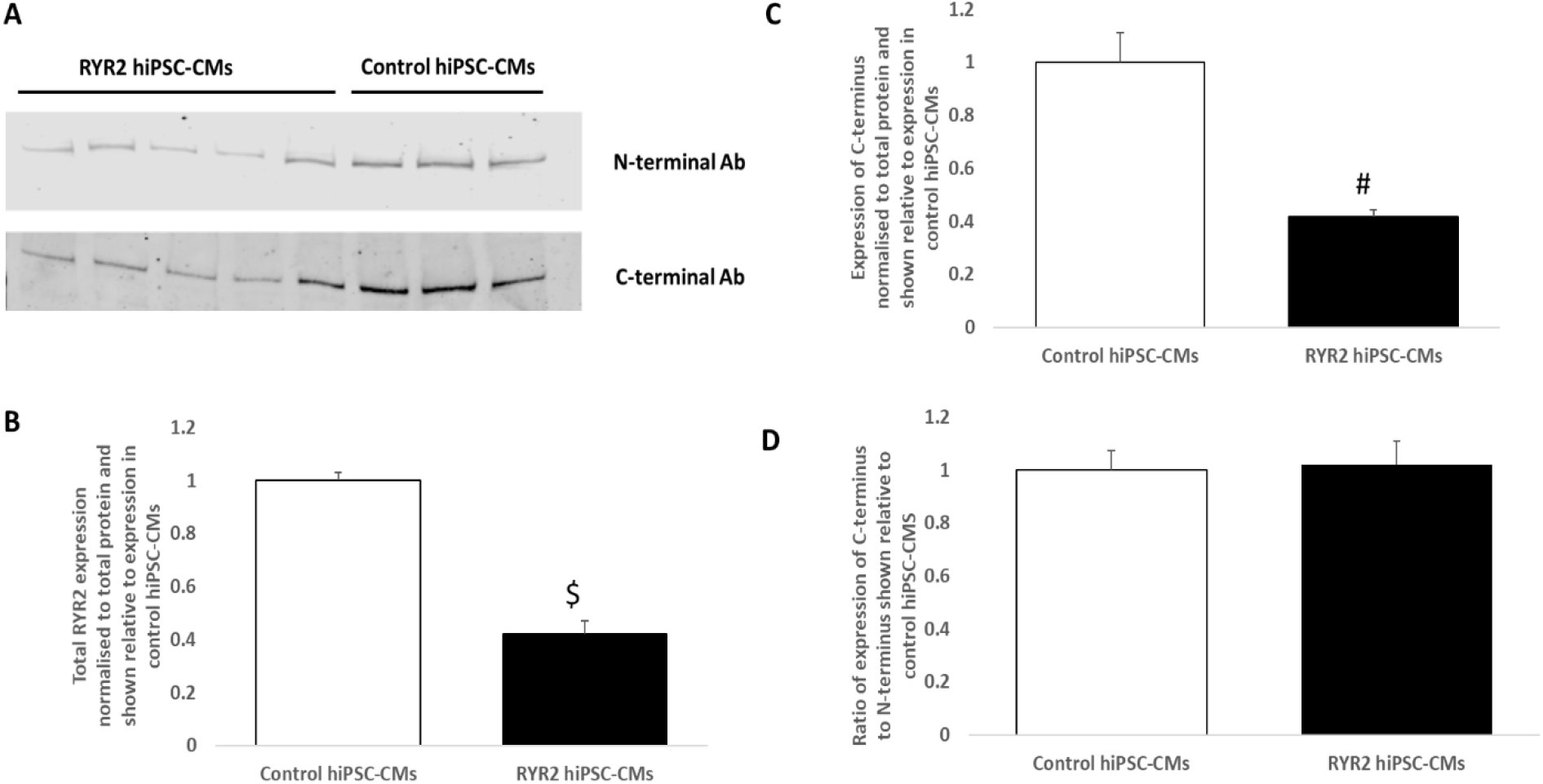
Western blot to assess RYR2 levels. (A) Representative western blots showing expression of N-terminus and C-terminus in protein extracted from RYR2-hiPSC-CMs and control hiPSC-CMs. (B) Quantification of expression of N-terminus (total) of RYR2 in RYR2 and control hiPSC-CMs. Data normalised to total protein. Total RYR2 expression was significantly reduced in the RYR2-hiPSC-CMs compared to the control hiPSC-CMs ($ p<0.0005). (C) Quantification of expression of C-terminus of RYR2 in RYR2 and control hiPSC-CMs. Data normalised to total protein. Expression of the C-terminus in RYR2 hiPSC-CMs was significantly reduced compared to expression in control hiPSC-CMs. # p<0.001. (D) Ratio of expression of C-terminus to N-terminus in RYR2-hiPSC-CMs shown relative to expression in control hiPSC-CMs. No significant difference was seen between the RYR2 and control hiPSC-CMs. n=5 (RYR2) and n = 3 (control).

### Partial silencing of the RYR2 allele harbouring the nonsense variant reduces calcium handling abnormalities

In order to ascertain whether the abnormal calcium handling phenotype seen in the RYR2-hiPSC-CMs is due to an effect caused by the presence of the p.(Arg4790Ter) RYR2 allele, allele-specific shRNAs were used to silence the mutant allele. The effect of seven different *RYR2* allele-specific shRNAs on the expression of the mutant *RYR2* allele in the RYR2-hiPSC-CMs was assessed. RT-PCR using allele-specific primers based on the rs684923 SNP in *RYR2* was performed. Segregation studies performed on family members of the proband identified that the T allele segregated with the nonsense variant and could be used to reflect expression of the mutant allele. shRNA_11 (in which the mutation recognition site was located at position 11 from the 5 ‘end of the shRNA) resulted in a significant reduction in expression of the mutant allele (Figure 7A) and a significant increase in the wild-type/mutant allele expression (Figure 7B).

**Figure 7.**
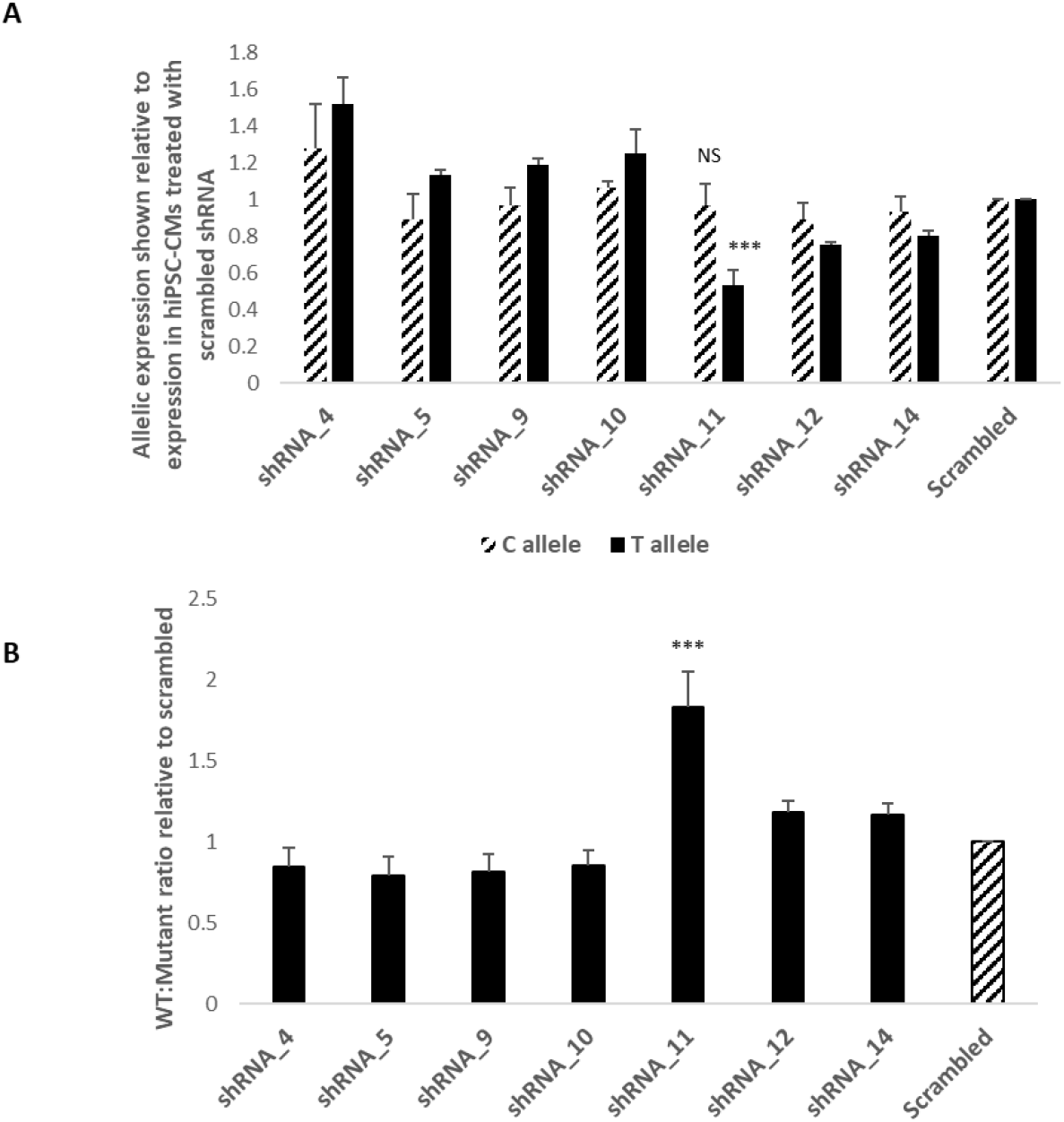
Allele-specific RT-PCR based on rs684923 performed on RNA extracted from RYR2-hiPSC-CMs treated with allele-specific shRNAs and a scrambled shRNA. (A) Allelic expression was normalised to total *RYR2* expression and is shown relative to the expression in the hiPSC-CMs treated with the scrambled shRNA. shRNA_11 led to a significant reduction in expression of the mutant allele (T allele) compared to treatment with the scrambled shRNA (*** p<0.005). There was no significant difference (NS) between expression of the wild-type (C allele) in the hiPSC-CMs treated with shRNA_11 compared to those treated with the scrambled shRNA. n=3 independent experiments. (B) The wild-type to mutant *RYR2* allelic ratio in hiPSC-CMs treated with allele-specific shRNAs, shown relative to scrambled. shRNA_11 results in the largest increase in wild-type:mutant ratio (*** p<0.005). n =3 independent experiments.

To determine the functional outcome of partial silencing of the mutated allele, we repeated the calcium imaging experiments on the RYR2-hiPSC-CMs transduced with allele-specific shRNA_11. As shown in the representative calcium transients and in the quantitative summary of the percentage of cells displaying calcium transient abnormalities (Figure 8), shRNA_11 treated RYR2-hiPSC-CMs displayed a significant reduction in calcium handling abnormalities compared to cells treated with the scrambled shRNA (40.5% vs 68.2%).

**Figure 8.**
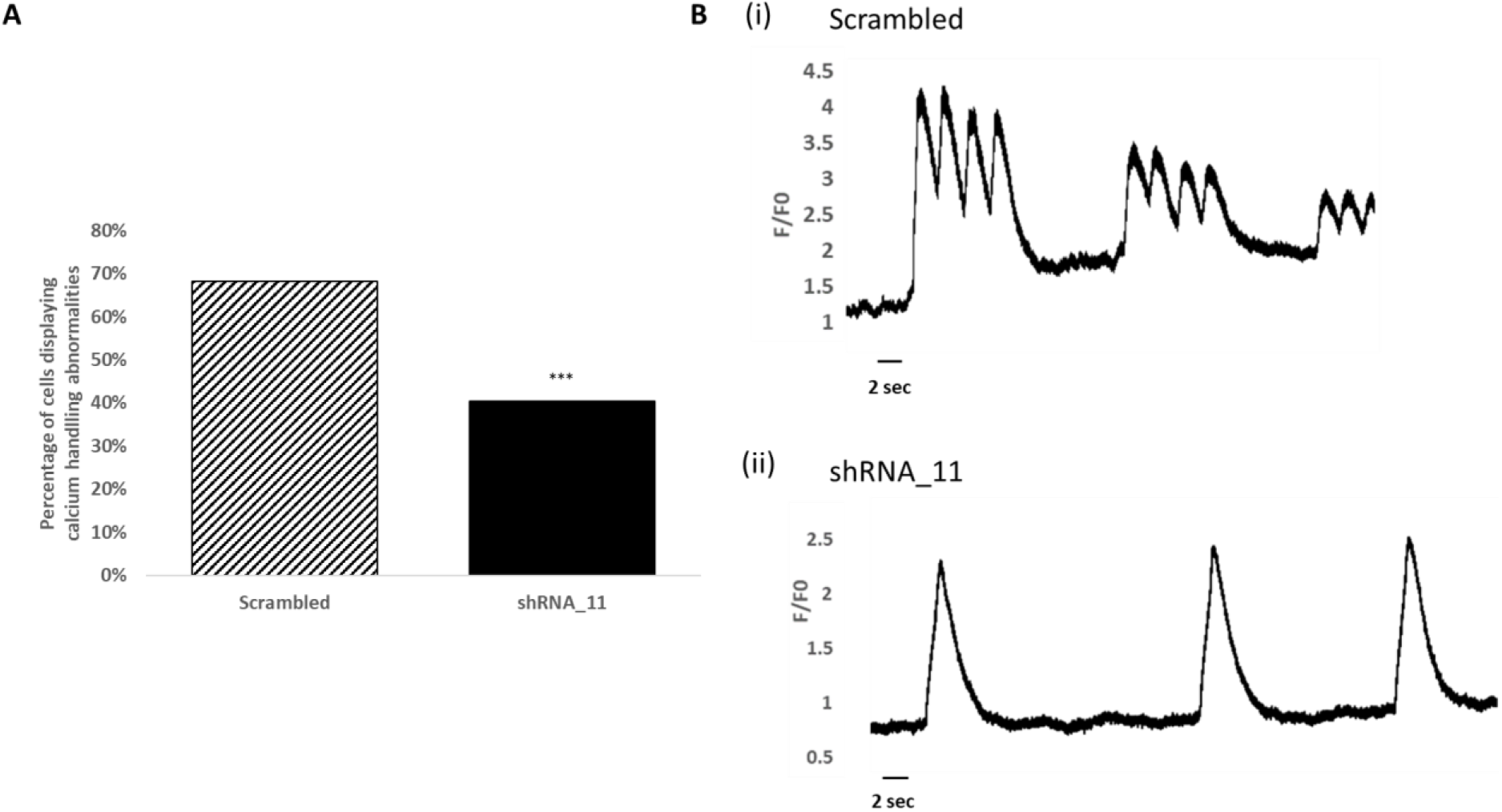
The effect of treatment with shRNA_11 on calcium handling in RYR2-hiPSC-CMs. (A) Treatment with shRNA_11 led to a significant reduction in the proportion of cells displaying abnormalities of the calcium transient compared to those treated with the scrambled shRNA. 68.2% of cells treated with scrambled (n=22) and 40.5% of cells treated with shRNA_11 (n=27) displayed calcium transient abnormalities (***p<0.005). (B) (i) Line scan image showing changes in intracellular calcium in a single RYR2-hiPSC-CM treated with the scrambled shRNA. (ii) Line scan image showing changes in intracellular calcium in a single RYR2-hiPSC-CM treated with shRNA_11.

## Discussion

In this study we have used hiPSC-CMs to elucidate the arrhythmogenic mechanism of a nonsense variant of the *RYR2* gene detected in a young patient who suffered a cardiac arrest. To the best of our knowledge this is the first described nonsense variant of *RYR2* associated with an arrhythmogenic phenotype. We have made two important observations; 1) The nonsense variant causes profound alterations in calcium handling that are likely to cause arrhythmias 2) These alterations are due to a gain of function of the cardiac ryanodine receptor and are abolished by inhibitors of RYR2.

### The p.(Arg4790Ter) variant causes profound calcium handling alterations that are likely to cause arrhythmias

The RYR2-hiPSC-CMs displayed significant calcium handling abnormalities characterised by prolonged transients with multiple peaks. These abnormalities can be triggered by isoproterenol. Similar abnormalities of the calcium transient have been previously reported in hiPSC-CMs derived from patients with CPVT caused by *RYR2* variants (3, 8, 9). In some of these studies it has been shown that these abnormalities lead to the formation of delayed afterdepolarizations (DADs) and phase 3 early afterdepolarizations (3, 8). Although the effect of the calcium handling abnormalities observed in the RYR2-hiPSC-CMs on the action potential was not assessed it is reasonable to hypothesize that the same mechanism occurs.

Although the presence of the truncated protein was not confirmed, treatment with the allele-specific shRNA, which caused a reduction in the expression of the mutant allele, resulted in a reduction in calcium handling abnormalities supporting the conclusion that expression of the p.(Arg4790Ter) allele is important in the development of the arrhythmogenic phenotype.

### Are the calcium handling abnormalities produced by the p.(Arg4790Ter) RYR2 variant due to a gain or a loss of function?

Most variants of *RYR2* known to cause CPVT result in a gain of function causing an increase in channel sensitivity to calcium facilitating the onset of spontaneous calcium release and DADs following adrenergic stimulation. It has been suggested that some *RYR2* variants can result in a loss of function due to a decreased sensitivity of the channel to activation by calcium (1, 10). Loss of function leads to an excessive accumulation of calcium in the sarcoplasmic reticulum that predisposes to early afterdepolarizations (1).

Three main lines of evidence support the conclusion that the p.(Arg4790Ter) variant acts in gain of function; 1) The calcium handling abnormalities observed in the RYR2-hiPSC-CMs at baseline were largely multiple peaked transients. These abnormalities have previously been reported in hiPSC-CMs harbouring gain of function variants of *RYR2* (3, 9). These abnormalities may represent calcium waves occurring during the calcium transient whilst the broader multiple peaked transients could potentially represent multiple calcium sparks. The RYR2-hiPSC-CMs also developed calcium handling abnormalities in response to treatment with isoproterenol. 2) The RYR2-hiPSC-CMs also displayed an increased tendency to develop spontaneous calcium release at various external calcium concentrations in comparison to the control hiPSC-CMs. This suggests a reduced threshold for SOICR, a key characteristic of gain of function *RYR2* variants. 3) The calcium handling abnormalities were largely abolished by treatment with nebivolol or carvedilol, both of which stabilize RYR2. Taken together these three lines of evidence strongly support the conclusion that the p.(Arg4790Ter) variant results in a gain of function.

### How does the p.(Arg4790Ter) variant lead to a gain of function?

The RT-PCR clearly demonstrated that the p.(Arg4790Ter) *RYR2* variant is expressed at mRNA level and escapes nonsense mediated decay. However, western blotting failed to confirm the presence of a truncated protein. A significant reduction in total RYR2 protein levels was observed. On the basis of these data we are not able to explain how the p.(Arg4790Ter) *RYR2* variant leads to a gain of function. We can only speculate and provide two hypotheses; 1) The mutant protein is undetectable but present at a level sufficient to disrupt channel gating and calcium handling; 2) The substantial reduction in total RYR2 levels caused by the variant results in remodeling of the calcium handling machinery that leads to a gain of function.

It is unclear how sensitive the western blotting approach using two different RYR2 antibodies is and whether it is able to detect low levels of the truncated protein. If translated, the truncated protein resulting from the p.(Arg4790Ter) variant would be missing part of the calcium sensing gate. It is plausible that incorporation of one truncated monomer into the channel could lead to significant alterations in calcium handling. In view of this it is reasonable to hypothesize that a low level of mutant protein could be sufficient to produce substantial disruption of calcium handling. Abnormal gating of half to a third of the total RYR2 tetramers would be sufficient to induce significant disruption of calcium handling similar to that observed in the RYR2-hiPSC-CMs. If we assume that between a third and a half of all RYR2 tetramers contain one truncated monomer the truncated monomers would represent between 8 and 12.5% of total RYR2. It is unclear whether the approach we utilized would enable us to detect these levels of expression.

Our data suggest that both *RYR2* alleles are expressed at mRNA level. However, the RYR2-hiPSC-CMs displayed a significant reduction in total RYR2 compared to the control hiPSC-CMs at both mRNA and protein level. To ascertain whether this could be due to cell line variation, expression of total *RYR2* was assessed in hiPSC-CMs derived from another independent control line. Expression of total *RYR2* in these control hiPSC-CMs was not significantly different from the expression in the control cells used in this study. It is possible that the variant leads to reduced RYR2 levels and the loss of RYR2 initiates a complex remodeling of the calcium handling machinery that leads to an overall gain of function. This is of particular interest if we consider that the *RYR2* gene is highly intolerant of haploinsufficiency. In the gnomAD database *RYR2* has a probability of loss of intolerance (pLI) of 1.0 (4). Further work is needed to assess the effects of reduction of RYR2 levels.

### Limitations

It is well understood that hiPSC-CMs do not display the structural organization seen in adult human cardiomyocytes however recently protocols have been developed to circumvent these issues including the development of T-tubules which allow the close apposition of L-type calcium channels with ryanodine receptors (11). Although there has been some data to suggest that hiPSC-CMs over 21 days of age display a robust calcium handling phenotype (12), clearly the development of a more mature structural phenotype would improve this. The hiPSC-CMs used in this study were all over 25 days old and it is possible that some of the calcium handling abnormalities observed at baseline may have been due to a degree of immaturity, however abnormalities due to this would be expected to be comparable between the RYR2 and control hiPSC-CMs.

One significant limitation of this study is the lack of an isogenic control line. This would have allowed the observed differences to be more directly attributed to the variant being studied. However, despite this, this work provides further insight into the mechanisms by which *RYR2* variants can lead to arrhythmias and also again highlights that hiPSC-CMs can be used as an effective tool to provide insights into disease mechanisms and testing potential therapeutic agents.

## Conclusion

The p.(Arg4790Ter) variant appears to behave like other gain of function variants in *RYR2*. The calcium handling abnormalities observed in the RYR2-hiPSC-CMs were improved with treatment with carvedilol and nebivolol suggesting these may be effective treatments for patients with gain of function variants in *RYR2*.

## Data Availability

Original source data and further clinical details can be provided upon request.

