## Supplementary Materials for "Characterisation of the mechanism by which a nonsense variant in *RYR2* results in ventricular arrhythmia"

***hiPSC generation***

A 3mm punch skin biopsy was obtained from which dermal fibroblasts were isolated and cultured. Fibroblasts were passaged using 0.25% Trypsin (Gibco).

HEK293GP cells were transfected with plasmids containing sequences encoding for human transcription factors (pMXs-hOCT4-Plath (Addgene plasmid #17964, (1)), pMXs-hSOX2 (Addgene plasmid #17218, (2)), pMXs-hKlf4 (Addgene plasmid #17219, (2)) along with a packaging plasmid VSVG using the transfection reagent PolyJet. A plasmid containing GFP was also used to allow visual confirmation of whether the transduction of the human dermal fibroblasts had been successful. Two days following the transfection, virus containing media was removed and filtered with a 0.45µm filter. The filtered media was then put into a centrifugal concentrator (Vivaspin) and was centrifuged at 6,000g for 20 minutes. 3µg/ml polybrene was then added to the concentrated virus containing media. This was added dropwise to dermal fibroblasts which had been passaged the day before. 72 hours following the initial transfection of the HEK293GP cells a second transduction of the dermal fibroblasts was performed. One week later the fibroblasts were manually passaged and plated onto mouse embryonic fibroblasts (MEFs). The cells were cultured in SR media (0.1mM beta-mercaptoethanol (Gibco), non-essential amino acids (Invitrogen), 1mM glutamine (Gibco), 20% knock-out serum replacement (Gibco), recombinant human FGF 10ng/ml (R&D Systems), 0.9mM valproic acid (Calbiochem)) and the media was refreshed daily. The cells were plated onto fresh MEFs every two weeks. Approximately 3 weeks following transduction hiPSC colonies were identified and manually picked and plated into 24 well tissue culture treated plates containing MEFs. The cells were cultured in SR media which was supplemented with 5µM Y27632 (Cayman Chemical), a Rock inhibitor, for the first two days.

hiPSC colonies were passaged using collagenase type IV (325units/ mg, Worthington). Cells were incubated with Collagenase IV for 40 minutes at 37$^{o}$C. The cells were then collected up in SR media and centrifuged twice at 100g for 4 minutes. The cell pellet was then resuspended in fresh SR media which had been supplemented with the rock inhibitor, Y27632 (5µM). The cell suspension was then plated into wells containing MEFs. When the colonies appeared larger they were passaged into tissue culture treated 6 well plates which were pre-coated with Growth Factor Reduced Matrigel (1:200, Corning).

Once established, hiPSCs were cultured in mTeSR1 culture medium (Stem Cell Technologies) on Matrigel. Karyotype analysis (Supplementary Data – Figure S2) and testing for Mycoplasma was regularly performed on the hiPSCs.

A well-established control hiPSC line generated using the same method was used in this study.

***Differentiation of hiPSCs into cardiomyocytes***

When the hiPSCs reached approximately 80% confluence the differentiation protocol was initiated. Cells were cultured in CDM3 medium (RPMI 1640 supplemented with recombinant human albumin (Sigma), L-ascorbic acid (Sigma) and 1% penicillin streptomycin (Sigma)) supplemented with 6µM CHIR99021 (Sigma) for two days after which the medium was changed and the CDM3 was supplemented with 2µM Wnt-C59 (Selleck Chemicals). Two days later the medium was changed to a medium (RPMI-B27 medium) consisting of RPMI 1640, 2% B27 supplement minus insulin (Life Technologies) and 1% penicillin streptomycin. The media was refreshed every other day until spontaneously beating cells were seen at around 10 days.

Two hiPSC clones were used in experiments, RYR2-1 and RYR2-3. Once the pluripotency and their ability to differentiate into hiPSC-CMs was established and no significant differences in baseline calcium handling was observed between the two clones the data from these two clones was pooled for subsequent experiments.

Prior to RNA and protein extraction hiPSC-CMs were cultured in CDM3 media containing lactate and no glucose. This allowed the cells to be purified ensuring that the only cells remaining were cardiomyocytes. This media was made up of RPMI 1640 minus glucose, L-ascorbic acid, human albumin, penicillin and streptomycin and lactate. The cells were treated with lactate containing media for 7 days starting on day 10 of the differentiation protocol.

***Immunostaining***

Undifferentiated cells were fixed with 4% (w/v) paraformaldehyde (Alfa Aesar). 10% (v/v) donkey or goat serum was used for blocking. For nuclear and cytoplasmic staining 0.1% Triton X-100 (Sigma) was included in the blocking solution. Following washes with D-PBS, primary antibodies for the pluripotency markers OCT4 (1:100, BD), SSEA4 (1:200, R&D Systems), TRA-1-60 (1:200, Abcam) and NANOG XP (1:400, NEB) were applied and left to incubate overnight at 4°C. Samples were washed and then incubated with secondary antibodies for one hour at room temperature. Nuclei were stained with DAPI (1:5000, Invitrogen). The specimens were then examined using an Olympus inverted microscope.

Cells which had been differentiated into cardiomyocytes were stained for cardiac specific markers. The cells were fixed, permeabilised and blocked as described above. Primary antibodies for the cardiac specific markers, alpha-actinin (1:150, Sigma Aldrich, A7811) and troponin I (1:150, Merck, MAB1691) were applied and the preparations were incubated overnight at 4$^{o}$C. Following this, the preparations were washed with D-PBS and then incubated with appropriate secondary antibodies for one hour at room temperature. Nuclei were stained with DAPI (1:500, Invitrogen). The cells were then visualised using a laser confocal microscope (Zeiss LSM-710).

***RNA extraction and RT-PCR***

RNA was extracted from cells using TRIzol (Life Technologies) according to manufacturer's protocol. DNase treatment was carried out on 750ng RNA. Reverse transcription into cDNA was undertaken using Superscript II (Invitrogen).

Real-time quantitative PCR (qPCR) was performed in duplo using SYBR green (Roche) in the StepOne plus Real-Time PCR machine (Applied Biosystems). Conditions were as follows; 5 mins at 95$^{o}$C, 40 cycles of 10 seconds at 95$^{o}$C, 20 seconds at 60$^{o}$C, 20 seconds at 72$^{o}$C followed by a step and hold meltcurve.

For allele-specific RT-PCR, allele-specific primers were designed on single nucleotide polymorphisms (SNPs) in *RYR2* and allelic expression was normalised to total *RYR2* expression.

The RT-PCR data was analysed using LinReg, a customised programme designed to analyse quantitative PCR data (3).

***Protein extraction and Western blot***

Protein was extracted from hiPSC-CMs. This was done by washing the cells with ice cold D-PBS and then scraping cells off from the bottom of the 6 well plate and resuspending in ice cold RIPA buffer containing protease and phosphatase inhibitors (0.1mg/ml phenylmethylsulfonyl fluoride (PMSF), 1mM sodium orthovanadate, 1ug/ml aprotinin, 1ug/ml leupeptin). The suspension was then kept on ice and intermittently vortexed for 30 minutes. The samples were then centrifuged for 30 minutes at 12,000rpm at 4$^{o}$C. The supernatant was aspirated and the pellet was discarded.

Protein quantification was undertaken using the Millipore Direct Detect system.

40µg of protein was loaded into each well of a 10 well NuPAGE 3-8% Tris-Acetate gel. 5µl HiMark pre-stained protein standard (Invitrogen) was loaded in one well of the gel. Prior to loading, the samples were incubated for 20 minutes at room temperature. The gel was run on ice at 150V for 2 hours followed by a further 40 minutes at 200V. The proteins were transferred onto a nitrocellulose membrane (GE Healthcare) using 20V for 16 hours on ice at 4°C. Following transfer, the membrane was stained with Revert Total Protein Stain (Licor) for 5 minutes at room temperature after which it was imaged with the Licor Odyssey Imaging System and then washed with TBS-T. The membrane was then blocked with 5% (w/v) non-fat dried milk solution for 1 hour at room temperature. After blocking the membrane was washed with TBS-T for 5 minutes three times. The membrane was then incubated with primary antibodies for 2 hours at room temperature and then washed with TBS-T for 5 minutes, three times. The membrane was then incubated with appropriate secondary antibodies (goat anti-mouse and goat anti-rabbit, both 1:10,000, Licor) for 1 hour at room temperature. Following incubation with the secondary antibodies the membrane was washed with TBS-T for 5 minutes three times. Protein bands were detected and densitometry analysis was performed using the Licor Odyssey Imaging System. The background signal was subtracted from all values and the lane normalisation factor was calculated by dividing the total protein signal for each sample by the largest total protein signal on the membrane. The densitometry values for protein bands were then divided by the corresponding lane normalisation factor producing a normalised signal value.

A ratio of the expression of the N-terminal antibody to the C-terminal antibody in the RYR2-hiPSC-CMs was calculated and then compared to the same ratio in the control hiPSC-CMs.

***Calcium imaging of hiPSC-CMs***

To prepare cells for imaging, cells were dissociated into Matrigel coated 35mm MatTek glass bottom dishes (MatTek Corporation). This was done by removing the medium and washing the hiPSC-CMs with 1ml D-PBS without Ca2+ or Mg2+. 1ml TrypLE Express was placed in each well and incubated at 37°C for 10 minutes. The cells were then removed from the bottom of the well and collected into 9ml of RPMI B27 medium. The medium containing the cells was passed through a 40µm cell strainer (ThermoFisher) and was centrifuged at 1,100 rpm for 5 minutes. The supernatant was discarded and the cell pellet was resuspended in 3ml of RPMI-B27 medium supplemented with 2µM thiazovivin. Varying amounts (75µl - 250µl) of the cell suspension were plated onto pre-prepared Matrigel coated MatTek glass bottom dishes. After 24 hours, a further 2ml of RPMI-B27 medium was placed onto cells in the MatTek dishes.

Between five and 10 days post dissociation, cells were loaded with 5µM Fluo-4 AM (Invitrogen). The cells were incubated with the medium containing Fluo-4 AM for 30 minutes at 37°C. This was then removed and the cells were washed with tyrodes solution and left to de-esterify for 30 minutes in normal tyrodes solution at 37°C before experiments were undertaken. The hiPSC-CMs used in calcium imaging experiments were between 25 and 70 days old from the start of differentiation. All imaging experiments were conducted at 37°C in tyrodes solution; (in mmol/l) NaCl 140, KCl 5.4, $CaCl_2$ 1.8, $MgCl_2$ 1 and glucose 10. In all experiments one abnormal calcium transient or ectopic calcium release was regarded as the minimum finding for arrhythmia and n refers to the number of cells.

***shRNA design and generation***

Allele-specific short hairpin RNAs (shRNAs) to target the mutant *RYR2* allele were designed. The allele-specificity was based on the nonsense variant in *RYR2* and in each shRNA the mutation recognition site was located in a different position (for shRNA sequences see Table S3). The shRNAs were cloned into a lentiviral vector containing a puromycin resistance cassette (plKO.1_puro).

4µg of the plKO.1_puro plasmid containing the shRNA was combined with packaging plasmids, pMDLg/pRRE, Addgene plasmid #12251 (4), pCMV-VSVg, Addgene plasmid # 8454, (5), and pRSV-Rev, Addgene plasmid #12253 (4), 2.7µg, 1.4µg and 1.0µg respectively (see Appendix for plasmid sequence maps). All plasmids were kindly donated by Prof Pinto's laboratory, AMC, The Netherlands. Transfection of HEK293T cells was performed using genejammer (Agilent). The following day, the culture media was removed and replaced with cardiomyocyte differentiation media (CDM3). The next day, the media was filtered using a 0.45µm filter and then added to the hiPSC-CMs.

The day after transduction, the media on the hiPSC-CMs was refreshed. Five days after transduction, the media on the hiPSC-CMs was changed and supplemented with puromycin (BioVision). Two days later the cells were washed with D-PBS and RNA was extracted using TrIzol. Expression of the mutant and wild-type alleles was then assessed using allele-specific RT-PCR.

For calcium imaging, the shRNAs were cloned into the plKO plasmid where puromycin was replaced by DsRed. hiPSC-CMs were transduced with virus produced using plasmids containing Ds-Red and either the allele-specific shRNA or a scrambled negative control shRNA. 20,000TU of virus were added to each MatTek plate. Between 5 to 7 days after transduction calcium imaging was performed on the cells. The cells were loaded with the fluorescent calcium indicator Fluo-4-AM. Cells which had been successfully transduced were identified by imaging using an evolve EMCCD camera and Micromanager software. After a cell had been identified as expressing DsRed, calcium imaging was performed as previously described.

***Genomic DNA extraction and sequencing***

Genomic DNA was extracted using the Isolate II Genomic DNA Kit (Bioline). Fragments of the *RYR2* gene were amplified by PCR using 100ng DNA and custom designed primers (see Table S1 and S2). Gel electrophoresis was used to confirm amplification of correct products and products were then isolated using the Isolate II PCR and Gel Kit (Bioline). Sanger sequencing was performed on all samples.

***Exome Sequencing***

Whole exome sequencing was performed on genomic DNA from proband. Enrichment was performed using the SureSelect Human All Exon Kit (version 5; Agilent) for the Illumina HiSeq 2500 system and variant analysis was undertaken using the Genome Analysis Tool Kit software (https://www.broadinstitute.org/gatk/). Variants with an allele frequency of greater than 0.005 were deemed common and were filtered out. The remaining variants were then compared to databases and in-house allele frequencies. *In silic*o prediction tools such as SIFT (6) and PolyPhen (7) were used to determine the likely pathogenicity of the variants. No likely pathogenic variants responsible for the phenotype in the proband were identified.

***Statistical Analysis***

Continuous variables were expressed as mean $\pm$ SEM and differences were assessed using the student's t-test. Categorical differences between groups were assessed using the chi-square test. A p value of less than 0.05 was deemed statistically significant.

| Target region / SNP | Forward Primer | Reverse Primer |
|---|---|---|
| Exon 100 *RYR2* | GCTTCTAGTAAACACGGCTGT | TCTTTAAACAGCCCTGCAACT |
| rs3765097 | ACAGTGATGTAGGGAGAGAG | TCAGGGCTCGTAGTCTGTTC |
| rs684923 | TCCCAGCGTCAAGCATGATG | AGATTCAGGTCCTTGGCTG |

**Table S1. Primers to check the presence of the p.(Arg4790Ter) variant and also two SNPs in *RYR2*, rs3765097 and rs684923.**

| Primer Name | Primer Sequence | Annealing Temperature ($^{o}$C) |
|---|---|---|
| RYR2_total_Fwd | ACAGAGTTTGGCACACAGCAG | 60 |
| RYR2_total_Rv | ACAGCAACATGACCACCATATCC | 60 |
| GAPDH_Fwd | ACCCACTCCTCCACCTTTGAC | 60 |
| GAPDH_Rv | ACCCTGTTGCTGTAGCCAAATT | 60 |
| RYR2_rs3765097Gen_Rv | TCTTCAGGGCTCGTAGTCTG | 66 |
| RYR2_rs3765097C_Fwd | TGCCTATAGAGTCCGTAAGC | 66 |
| RYR2_rs3765097T_fwd | TGCCTATAGAGTCCGTAAGT | 66 |
| RYR2_rs3765097Gen_Fwd | GATTTGCCTATAGAGTCCGTAAG | 66 |
| RYR2_rs684923C_Rv | TTAAGAGGCATCTTTGCG | 60 |
| RYR2_rs684923T_Rv | TTAAGAGGCATCTTTGCA | 60 |

| | | |
|---|---|---|
| RYR2_rs684923Gen_Fwd | TCCATAGAAGTTTGTTTACTCTC | 60 |
| RYR2_rs684923Gen_Rv | TTTAAGAGGCATCTTTGC | 60 |
| Nanog_Fwd | AGAATAGCAATGGTGTGACGCAG | 60 |
| Nanog_Rv | TGGATGTTCTGGGTCTGGTTGC | 60 |
| OCT4_Fwd | TGGTTGGAGGGAAGGTGAAG | 60 |
| OCT4_Rv | TGTCTATCTACTGTGTCCCAG | 60 |
| SOX2_Fwd | ACCAATCCCATCCACACTCAC | 63 |
| SOX2_Rv | TCTATACAAGGTCCATTCCCCC | 63 |
| MYH6_Fwd | TACAGGACCTGGTGGACAAGC | 60 |
| MYH6_Rv | TTGCGGAACTTGGACAGGTTG | 60 |
| cTNI_Fwd | AGTCACCAAGAACATCACGGAGAT | 60 |
| cTNI_Rv | GCAGCGCCTGCATCATG | 60 |
| MYH7_Fwd | AGAGTCGGTGAAGGGCATGAG | 60 |
| MYH7_Rv | AGCTTGTCTACCAGGTCCTG | 60 |

**Table S2. Primer sequences and annealing temperatures used in RT-PCR reactions**

| shRNA | Forward | Reverse |
|---|---|---|
| shRNA4 | ccggaaGCATTCAATTTTTTCTGAAtcaagacTTCAGAAAAAATTGAATGCtttttttg | aattcaaaaaaaGCATTCAATTTTTTCTGAAgtcttgaTTCAGAAAAAATTGAATGCtt |
| shRNA5 | ccggaaCATTCAATTTTTTCTGAAAtcaagacTTTCAGAAAAAATTGAATGtttttttg | aattcaaaaaaaCATTCAATTTTTTCTGAAAgtcttgaTTTCAGAAAAAATTGAATGtt |
| shRNA9 | ccggaaCAATTTTTTCTGAAAATTCtcaagacGAATTTTCAGAAAAAATTGtttttttg | aattcaaaaaaaCAATTTTTTCTGAAAATTCgtcttgaGAATTTTCAGAAAAAATTGtt |
| shRNA10 | ccggaaAATTTTTTCTGAAAATTCTtcaagacAGAATTTTCAGAAAAAATTtttttttg | aattcaaaaaaaAATTTTTTCTGAAAATTCTgtcttgaAGAATTTTCAGAAAAAATTtt |
| shRNA11 | ccggaaATTTTTTCTGAAAATTCTAtcaagacTAGAATTTTCAGAAAAAATtttttttg | aattcaaaaaaaATTTTTTCTGAAAATTCTAgtcttgaTAGAATTTTCAGAAAAAATtt |
| shRNA12 | ccggaaTTTTTTCTGAAAATTCTACtcaagacGTAGAATTTTCAGAAAAAAtttttttg | aattcaaaaaaaTTTTTTCTGAAAATTCTACgtcttgaGTAGAATTTTCAGAAAAAAtt |
| shRNA14 | ccggaaTTTTCTGAAAATTCTACAAtcaagacTTGTAGAATTTTCAGAAAAtttttttg | aattcaaaaaaaTTTTCTGAAAATTCTACAAgtcttgaTTGTAGAATTTTCAGAAAAtt |
| Scrambled | ccggaaGAAATGTACTGCGTGGAGAtcaagacTCTCCACGCAGTACATTTCtttttttg | aattcaaaaaaaGAAATGTACTGCGTGGAGAgtcttgaTCTCCACGCAGTACATTTCtt |

**Table S3. Sequence of forward and reverse shRNA oligonucleotide sequences.** The targeting portion is indicated by uppercase font. These oligonucleotides were annealed and cloned into the plKO.1_puro and plKO.1_DsRed plasmids.
