## Supplementary Data for "Characterisation of the mechanism by which a nonsense variant in *RYR2* results in ventricular arrhythmia"

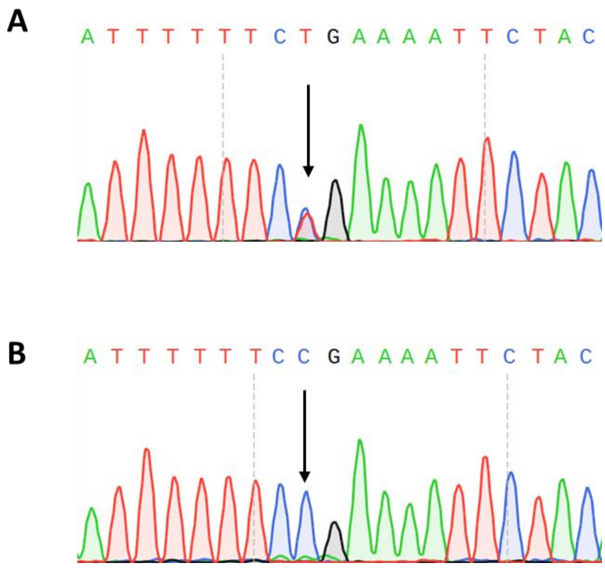


**Figure S1. Sequencing of exon 100 in *RYR2* in RYR2 and control hiPSCs.** (A) Sequencing of gDNA extracted from RYR2-1 hiPSCs confirmed the presence of the c.14368C>T p.(Arg4790Ter) variant. (B) Sequencing of gDNA from control hiPSCs confirmed the absence of this variant.

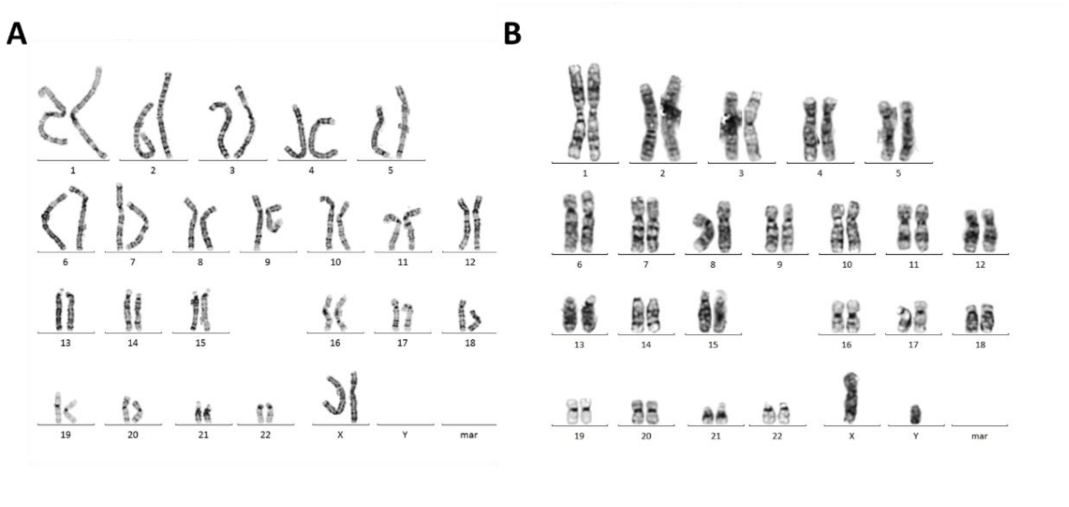


**Figure S2. Karyotypes of hiPSCs**. (A) Karyogram of RYR2-1 hiPSCs showing a normal female karyotype 46XX. (B) Karyogram of control hiPSCs showing a normal male karyotype, 46XY.

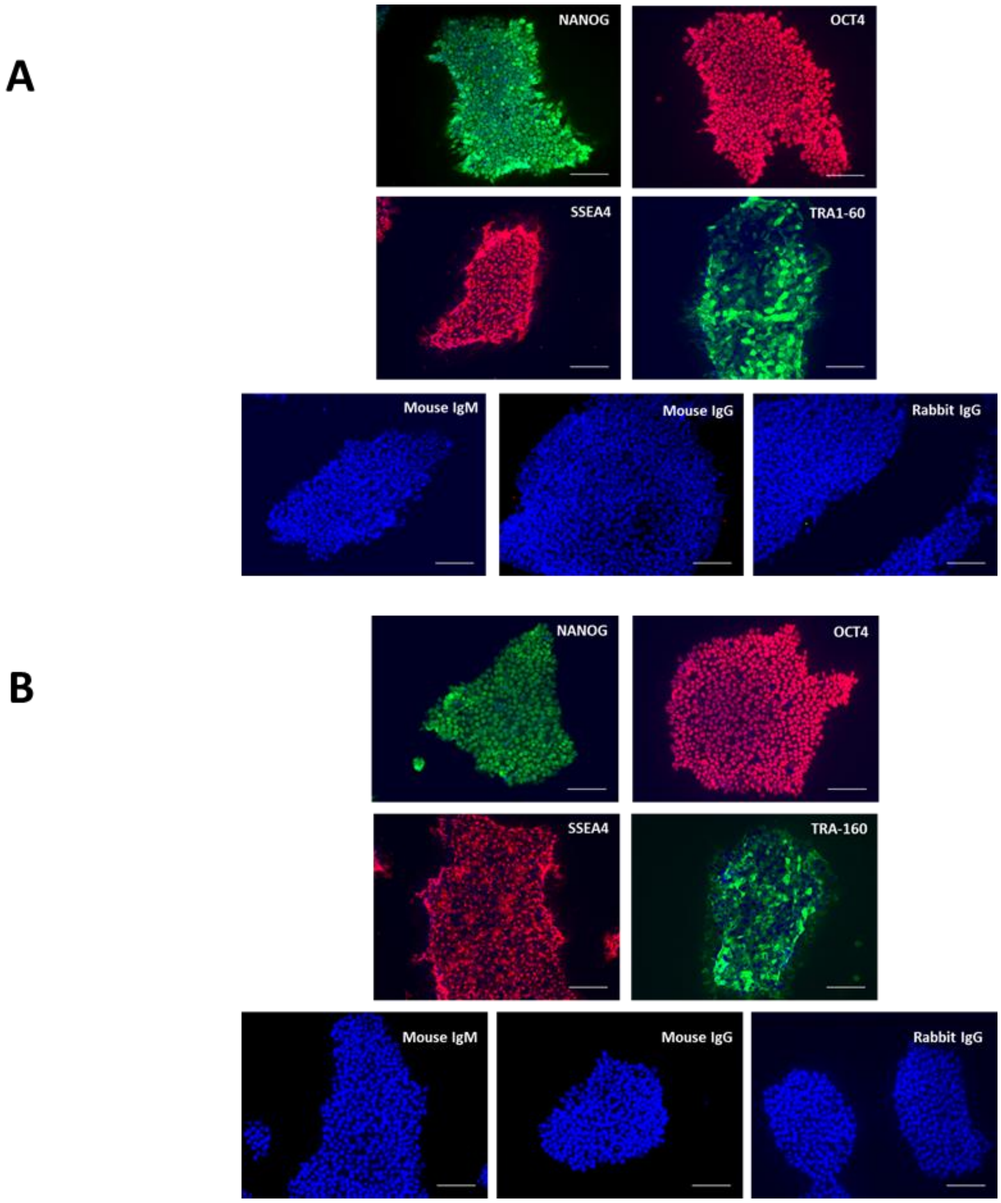


**Figure S3. Immunostaining of RYR2 and control hiPSCs for pluripotency markers.** Immunostaining of RYR2-1 (A) and control (B) hiPSCs for the pluripotency markers, Nanog, OCT4, SSEA4, TRA1-60 and also isotype controls (mouse IgM, mouse IgG and rabbit IgG). Nuclei stained with dapi (blue). Scale bars 100μm.

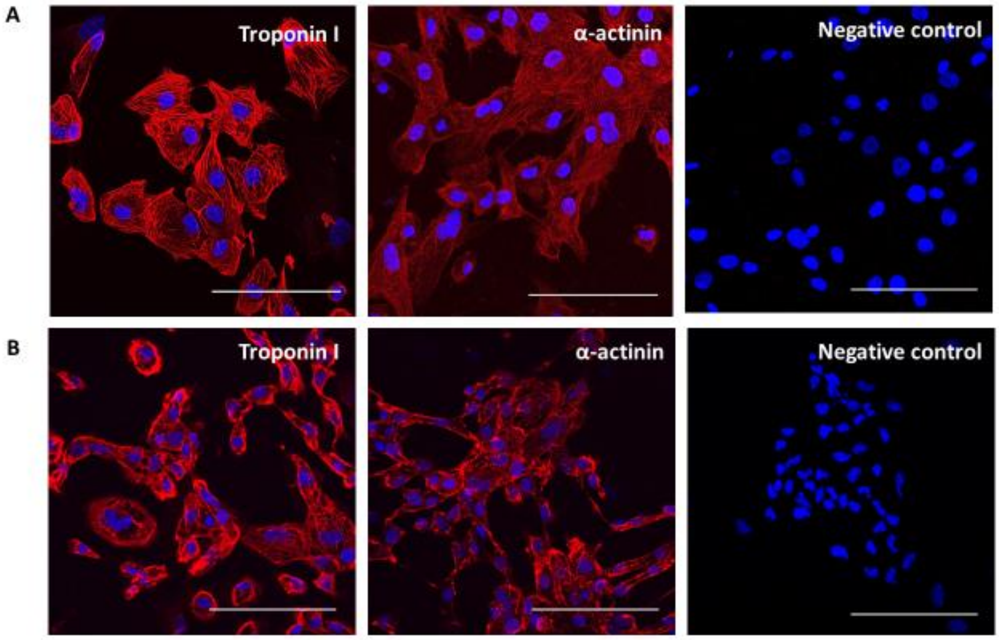


**Figure S4. Immunostaining of RYR2 and control hiPSC-CMs for cardiac markers.** Immunostaining of RYR2-1 (A) and control (B) hiPSC-CMs for the cardiac markers Troponin I (red) and α-actinin (red). Negative control (no primary antibody applied). Nuclei stained with dapi (blue). Scale bars 100µm.

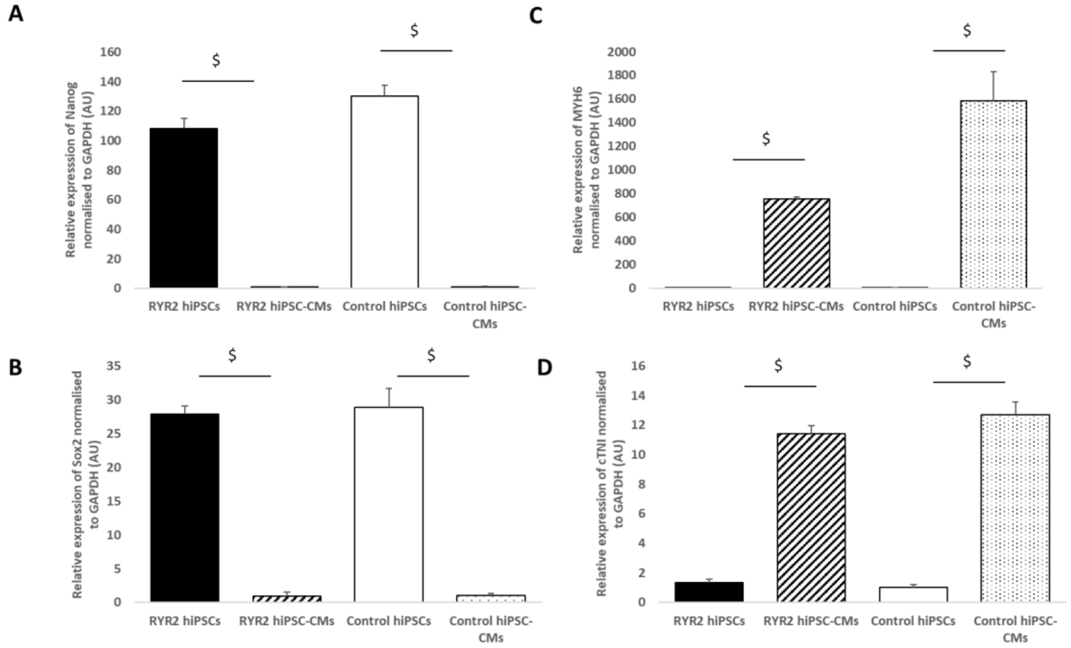


**Figure S5. RT-PCR to assess expression of pluripotency and cardiac markers in hiPSCs and hiPSC-CMs.** RT-PCR performed on cDNA synthesised from RNA extracted from RYR2-1 and control hiPSCs and hiPSC-CMs to assess the expression of Nanog (A), Sox2 (B), MYH6 (C) and cTNI (D) in RYR2 and control hiPSCs and hiPSC-CMs. Values normalised to GAPDH and expressed in arbitrary units (AU). Undifferentiated RYR2-1 and control hiPSCs displayed significantly higher expression of the pluripotency markers Nanog and Sox2 compared to the differentiated cells whilst the RYR2-1 and control hiPSC-CMs expressed significantly higher levels of the cardiac markers MYH6 and cTNI compared to the undifferentiated cells ($ $p<0.0005$). n=3 each group. Errors bars represent SEM.
